# Exploring Patterns and Trends in COVID-19 Exports from China, Italy, and Iran

**DOI:** 10.1101/2020.09.09.20190983

**Authors:** M.L. McHenry, A. Soliman, B. Dailey, T. Chen, J.J. Letterio, G. Luo

**Affiliations:** Case-Western Reserve University School of Medicine, Department of Population and Quantitative Health Sciences; Case-Western Reserve University School of Medicine, Department of Pediatrics; Case-Western Reserve University School of Medicine, Case Comprehensive Cancer Center; Duke University, Trinity College of Arts and Sciences; The Angie Fowler Adolescent & Young Adult Cancer Institute, University Hospitals Rainbow Babies & Children’s Hospital.

## Abstract

This paper investigates COVID-19 exported cases by country and the time it takes between entry until case confirmation for the exported cases using publicly available data. We report that the average days from entry to confirmation is 7.7, 5.0 and 4.7 days for travelers from China, Italy, and Iran respectively. Approximately, one-third of all exported cases were confirmed within 3 days of entry suggesting these travelers were mildly symptomatic or symptomatic in arrival. We also found that earlier exported cases from each country had a longer time between entry to confirmation by an average of 3 days compared to later exports. Based upon our data, reported exported cases from South Korea were far fewer in comparison to those from China, Italy and Iran. Therefore, we suggest that careful monitoring of likely symptomatic travelers and better public awareness may lead to faster confirmation as well as reduced transmission of COVID-19 pandemic.

## Introduction

Coronavirus Disease 2019 (COVID-19) originated in Wuhan, China, where cases were first reported in December 2019. The virus spreads through human to human transmission, leading to transmission in other countries by international travelers [1, 2]. Although China instituted a lockdown on Wuhan on January 23^rd^, 2020, more than 100 exported cases from China were reported by February 14^th^, 2020 [3, 4]. The Wuhan travel ban had a significant reduction on international exportation of COVID-19, but there was still undetected community spread outside of China [4]. In February 2020, large outbreaks of local transmission of COVID-19 were reported in South Korea, Italy, and Iran [5]. We decided to examine these three countries because they experienced some of the largest outbreaks in the time immediately after the initial outbreak in China and thus provide important insight into the spread of the virus through travelers from Wuhan. Unfortunately, we could only gather enough data from Italy, Iran and China, so these are the main focus of the paper. By mid-March, over 100 countries had reported cases of COVID-19 in their countries, and the World Health Organization (WHO) declared COVID-19 as a pandemic [6, 7]. Assuming that the virus did not arise independently in multiple locations, an outbreak of COVID-19 outside of China requires an infected traveler to bring the virus into the country and subsequently infect others. For countries without large quantities of local transmission of COVID-19, the priority is to prevent spread of the virus from infected travelers [8]. To achieve this, many countries have issued travel restrictions or travel bans for high risk countries (i.e. those with high numbers of COVID-19 cases) [9].

For our study, we investigated the exported cases from China, Italy, and Iran to observe when they traveled and when they were subsequently confirmed to have COVID-19. We used this data to observe the dates of exported cases, dates of confirmation of COVID-19, and time between entry to confirmation. Analyzing this data provided us insights into understanding how to better prevent and monitor international spread of the COVID-19.

## Methods

### Data Collection

Exported COVID-19 cases were collected from January 2020 to March 20^th^, 2020. We used a previously published study by Chinazzi et al. for exported cases from China (**Supplementary Table 1**) [10]. For cases exported from Italy and Iran, the information was gathered through government official statements, credible reports from foreign ministries of health focusing on the COVID-19 infection as well as published news articles from several agencies available online for the public (**Supplementary Table 2**). Most cases were ascertained from news articles but some were ascertained through government health reports and others through peer-reviewed journal articles [11]. We organized infected cases that traveled in a group as a single cluster because this is how they were originally reported and it was not possible to parse out dates for an individual case within a source of data. As such, the dates of entry and confirmation are the same for each case within a cluster. Clusters ranged in size from 1-17. However, most clusters consisted of solely 1 case (**Supplementary Table 1, Supplementary Table 2**). The average cluster sizes for China, Italy, and Iran were 1.2, 1.2, and 1.9, respectively. Our gathered data about each cluster of COVID-19 cases included the following: date of entry of the travelers to their destination country, date of confirmation of COVID-19 of the case, and the estimated size of the affected group. If multiple people were affected, the first confirmation date was used. If no date of entry was listed, we excluded the cluster from other data such as entry date and time between entry and confirmation. Time-to-diagnosis of suspected cases based on the former data was calculated and the references for each were noted in the accompanying Supplemental Tables.

### Analysis of Data

We made graphs for dates of confirmation, date of arrival, and constructed a table to show the days between entry and confirmation for clusters of exported cases of each country (**Figure 1, Figure 2, Figure 3**). For early and late exports, we selected a time period of between 10 to 14 days from the first reported entry as early. Any case after the cutoff date was considered late. We then compared the mean time from entry to confirmation between earlier and later discovered cases of COVID-19 within each country using a student’s t-test. For comparison of time from entry to confirmation between exported cases from all the different countries, we used ANOVA to calculate a p-value.

**Figure 1.**
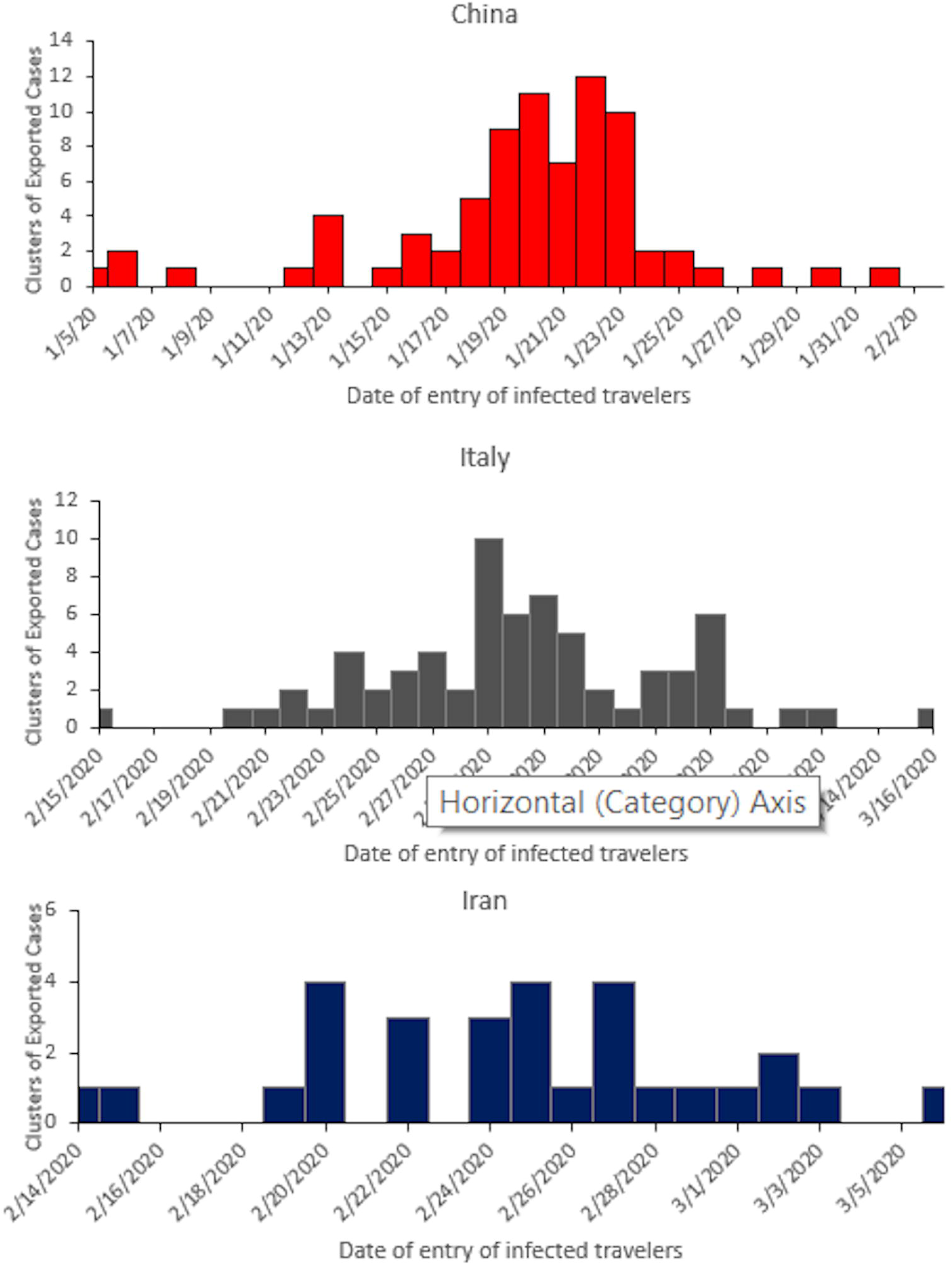
Exported cases by country according to entry date. Number of reported exported cluster of cases from A. China B. Italy C. Iran by date of arrival into foreign country.

**Figure 2.**
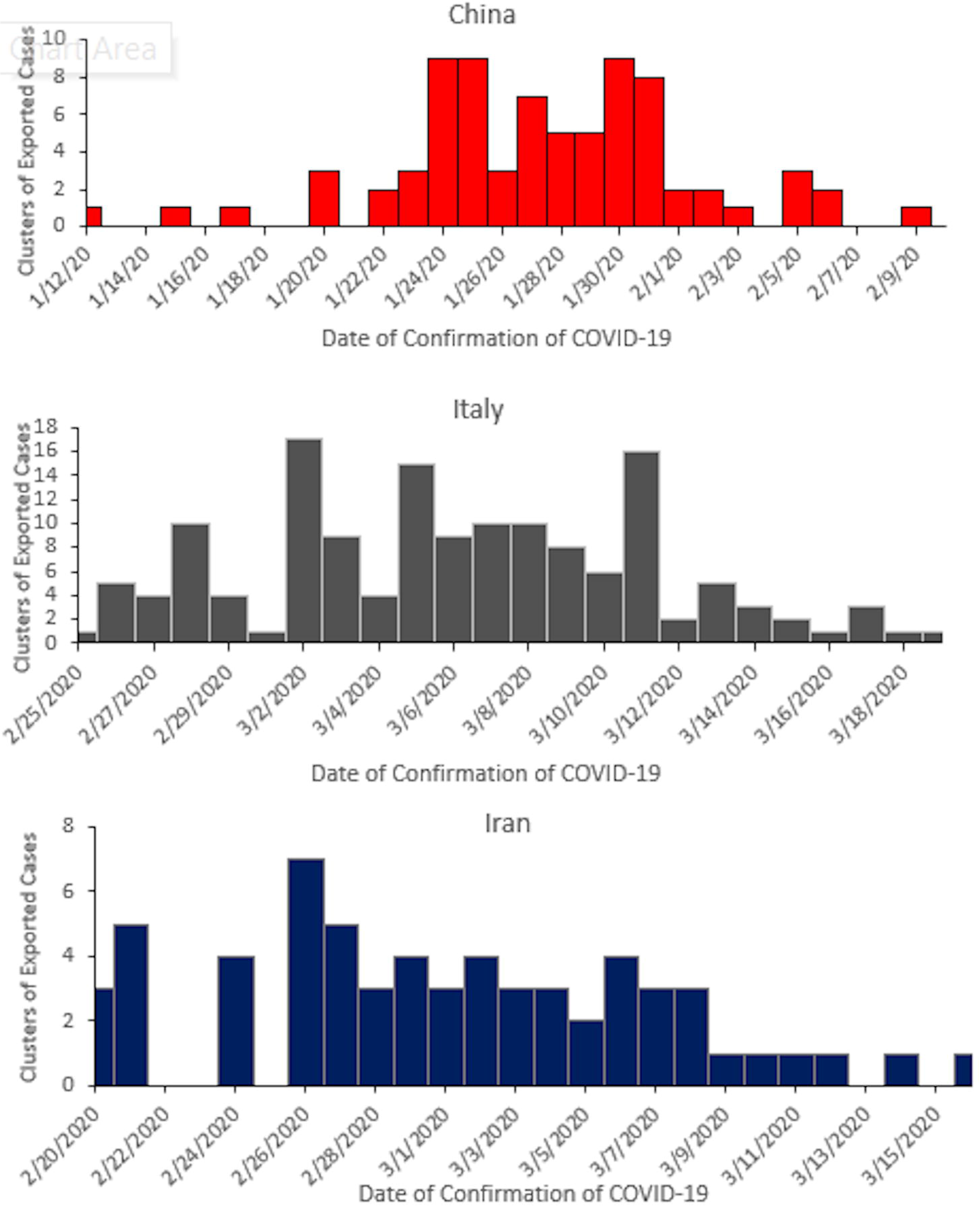
Exported cases by country according to Confirmation date. Number of reported exported cluster of cases from A. China B. Italy C. Iran by date of confirmation of COVID-19 into foreign country.

**Figure 3.**
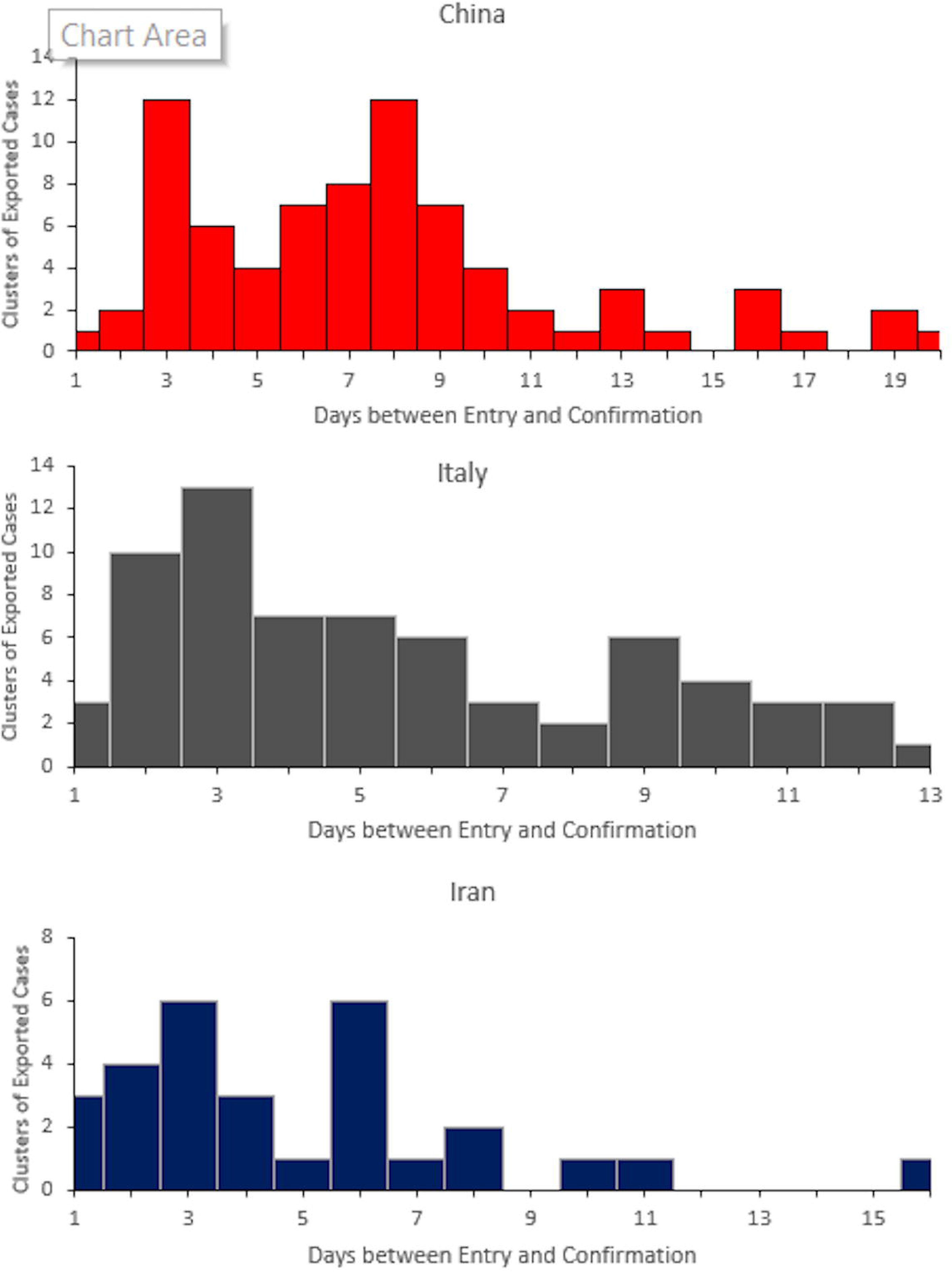
Exported cases by country according to days between Entry and Confirmation. Number of reported exported duster of cases from A. China B. Italy C. Iran by time between entry and confirmation of COVID-19 into foreign country.

## Results

Figure 1 shows the date of entry for exports for China, Italy, and Iran. COVID-19 infected cases left China as early as January 5th, 2020 (**Fig 1A**). Increased clusters of infected cases were noted between January 19th, 2020 and January 23rd, 2020 which dramatically decreased afterwards due to the Wuhan travel ban [4]. Infected cases from Italy entered other countries as early as February 15th, 2020 (**Fig 1B**). Subsequently, infected travelers increased to reach their highest level on February 29th, 2020. Due to the Italy travel ban announced on March 9th, 2020, a decrease in COVID-19 travelers is seen afterwards [12]. From Iran, infected travelers were entering other countries and transmitting the disease at a stable rate over the period from February 14th, 2020 to March 6th, 2020. Cases began to increase from February 20th until February 27th, 2020 (**Fig 1C**).

Figure 2 shows the clusters of COVID-19 exported cases from China, Italy and Iran according to their dates of confirmation. Exported cases from China were confirmed starting from January 12th, 2020 to reach two peaks on January 24th, 2020 and February 1st, 2020 before declining (**Fig 2A**). Interestingly, these two dates are the day after the Wuhan travel ban and approximately one week after the travel ban, respectively. Exported cases from Italy were confirmed starting on February 25^th^, 2020 and continued as they reached high numbers in early March 2020 (**Fig 2B**). Exported cases from Italy declined afterwards, likely due to implemented travel restrictions by the Italian government [12]. The confirmation of exported COVID-19 cases from Iran peaked on February 26th, 2020 then slowly declined since then (**Fig 2C**).

We then explored the approximate time between entry and confirmation of COVID-19 infection from China, Italy and Iran in foreign nations in days (**Figure 3**). For exported cases from China (**Fig 3A**), we observe a bimodal distribution with modes at 3 and 8 days between entry and confirmation which likely correlate with symptomatic (3 days) and asymptomatic cases (8 days) on arrival. Likewise, there are similar peaks at 3 days in exports from Italy (**Fig 3B**) and Iran (**Fig 3C**) suggesting these are cases of symptomatic or near symptomatic infected individuals. We calculate the mean time between entry and confirmation to be 7.7, 5.5, and 4.7 days for exports from China, Italy, and Iran respectively (**Table 1**). Exports from China had a significantly longer entry to confirmation time which is likely explained by the lack of testing kits at the start of this period (p < 0.05).

**Table 1.** Summary of Days from Entry to Confirmation

| Country | Dates of Entry (in 2020) | Clusters | Days from Entry to Confirmation: Mean (SD) | Student's t-test P-value* | ANOVA P-value□ |
| --- | --- | --- | --- | --- | --- |
| China | 1/5 – 2/1 | 77 | 7.7 (4.31) |  | <0.01 |
| Early China Exports | 1/5 – 1/19 | 29 | 9.5 (3.97) | <0.005 |  |
| Late China Exports | 1/20 – 2/1 | 48 | 6.6 (5.28) |  |  |
| Italy | 2/15 – 3/16 | 52 | 5 (3.28) |  |  |
| Early Italy Exports | 2/15 – 2/29 | 26 | 6.9 (3.30) | <0.001 |  |
| Late Italy Exports | 3/1 – 3/16 | 26 | 4 (2.63) |  |  |
| Iran | 2/14 – 3/6 | 28 | 4.7 (3.17) |  |  |
| Early Iran Exports | 2/14 – 2/26 | 21 | 5.5 (3.60) | ~0.010 |  |
| Late Iran Exports | 2/27 – 3/6 | 33 | 3.4 (1.75) |  |  |

\*A student's t-test was used to calculate p-values for the difference between early and late exports in mean days until COVID-19
□ Analysis of variance (ANOVA) testing was performed to calculate a p-value for the difference in days until confirmation between the three countries shown

We found that earlier exported cases (first 10-14 days after arrival of 1^st^ exported traveler) had a significantly longer time from entry to confirmation compared to later cases for each country (**Table 1**). For these three countries, the difference between earlier and later exports ranged from 2.1-2.9 days.

## Discussion

We performed a thorough search of government reports, news reports, and research articles for information about exported cases, but we had many cases without entry date while other cases had ambiguous information about the countries to which cases traveled (**Supplementary Table 3**). As a result we had to exclude these from our calculation of time between arrival and confirmation of COVID-19. While this is a notable limitation and it is possible that there are unmeasured differences between those we excluded and those in our analysis, we do not believe that their exclusion has biased our results, as these clusters had similar dates of confirmation and similar cluster sizes to those that were included (**Supplementary Table 4**). The only notable difference is that the cluster size was larger for exports from Italy with no date of entry, which was mostly due to a single cluster of 17 among the clusters with no date of entry. For China’s exported cases, the WHO tracked cases with travel history to China. Thus, our information for China is more complete and accurate [5]. However, our data did not include those who were evacuated due to COVID. While the inclusion of evacuees may have altered our results, people evacuated by their national government represent a different (likely lower) level of risk for transmission in their destination than those who travelled on their own. For Italy and Iran, we relied primarily on news outlets since there was no official tracking by any organization. Although our data is incomplete, we believe this study is the largest collection of current data on exports from Italy and Iran, two countries that initially had some of the highest burden of COVID-19 during the early stages of the pandemic and provide insight into how the virus was transmitted through at that time.

We attempted to ascertain exported cases from South Korea but were surprised at how few countries reported any (**Supplementary Table 3**). Due to such a small number of cases, we excluded this data from our calculation of time until confirmation but have included the clusters that we could identify. The very small number of exported cases from South Korea suggests that proper measures were taken to prevent spread of the virus to other countries. The very small number of exported cases from South Korea may suggest that proper measures were taken to prevent spread of the virus to other countries. These measures included early detection of local transmission and thorough testing of asymptomatic and suspicious contacts [13].

The time from entry to confirmation is a critical time in which travelers can spread the virus, potentially without being aware of doing so. Our estimated days from entry to confirmation is in line with the incubation period of approximately 5 days from previous studies [14]. The time between entry to confirmation is an important number for policymakers as it can help inform guidelines on travel restrictions, quarantine measures, and testing of individuals who have just entered the country. Further, it can help inform better epidemiological models of transmission The time from entry to confirmation for China’s exports was significantly longer compared to Italy and Iran. This is likely because testing for COVID-19 was still not readily available in many countries when exports first started and China was exporting cases earlier than Italy and Iran. There were even confirmed exported cases from China before January 10^th^, when the genome of the SARS-CoV-2 virus was first published [15, 16].

For all three countries, we observed that the earlier wave of exported cases had a longer time from entry to confirmation compared to later wave of cases from the same country (**Table 1**). It is unlikely that the incubation period of the virus would change during this time period; the shorter time until confirmation may be explained by more aggressive testing policies or an increased awareness of COVID-19 symptoms leading to earlier testing and confirmation. Early exports from Italy and Iran of COVID-19 were sometimes not tested for COVID-19 because these countries were not considered high-risk at the time [17]. It is also possible that earlier travelers were infected more immediately prior to their departure while later travelers were infected for many days before leaving. There are known examples of early exported cases experiencing symptoms for more than 3 days prior to confirmation of COVID-19 [18]. In our own data, 36.5% of all cases were confirmed in 3 days or less, indicating that they were likely symptomatic at the time that they left their country of exportation. Additionally, many COVID-19 cases, including exports from other countries, are detected through self-reporting to the hospital or clinic [19, 20]. This adds further weight to the argument that heightened awareness of COVID-19 in the later wave of exported cases likely contributed to the shorter time until confirmation observed in this group.

Two of the most common strategies to reduce transmission due to exportation is a complete travel ban or a 14-day quarantine of travelers from high-risk countries [9]. However, these measures are much more effective if they are implemented early. In the earliest stages of the pandemic, it was not immediately clear which countries were considered “high-risk” and by the time a country had reported outbreaks and enacted policies to reduce exportation of COVID-19 cases, there were already many exported cases. For example, South Korea implemented international travel reductions when they reported 200 COVID-19 cases domestically, and very few cases were exported as a result [21]. Other countries responded soon afterwards with travel recommendations or bans against South Korea, which reduced the number of exportations [22]. While Italian and Chinese lockdowns did help reduce exported cases significantly, there were already hundreds of exported cases at the time that these policies were enacted [4].This is illustrated by the high number of cases that were exported from China prior to the travel ban [3,4]. Similarly, as shown in this article, Iran and Italy exported many cases, which may have been a result of the delay in recognition of risk. This point is underscored by the fact that, in our data on these two countries, the later wave of exported cases had confirmed diagnoses much faster than the earlier ones, demonstrating that countries had a higher level of awareness and concern as time went on (**Table 1)**. In contrast, South Korea, which had vigilant surveillance and against which more timely travel bans were instituted, appears to have exported far fewer cases. Lastly, not all countries have the necessary resources or policies in place to adequately test for COVID-19, which may have greatly increased the undetected exported cases of COVID-19 [9].

Understanding the time until confirmation gives insight into how long people may be in the country and able to transmit the disease before being recognized as a COVID-19 case, a time during which they may have already begun to transmit the disease to others in their community. Further, our work shows how transmission continued to take place even as travel bans were implemented. As governments consider and implement travel bans during this pandemic, our research sheds light onto the risk posed by international travelers and provides valuable information on epidemiological surveillance of the spread of COVID-19 between countries. This information and analysis can lead to better epidemiological models, more informed decision making about how travel bans affect the exportation of cases, and how best to surveil and test travelers in order to minimize transmission of COVID-19.

## Data Availability

All data used to for the manuscript will be uploaded on medRxiv.

